# A gene pathogenicity tool ‘GenePy’ identifies missed biallelic diagnoses in the 100,000 Genomes Project

**DOI:** 10.1101/2023.03.21.23287545

**Authors:** Eleanor G. Seaby, Gary Leggatt, Guo Cheng, N. Simon Thomas, James J Ashton, Imogen Stafford, Genomics England Consortium, Diana Baralle, Heidi L. Rehm, Anne O’Donnell-Luria, Sarah Ennis

**Affiliations:** Human Development and Health, Faculty of Medicine, University Hospital Southampton, Southampton, Hampshire, SO16 6YD, UK; Program in Medical and Population Genetics, Broad Institute of MIT and Harvard, Cambridge, MA 02142, USA; Division of Genetics and Genomics, Boston Children’s Hospital, Boston, MA 02115, USA; Paediatric Infectious Diseases, Imperial College London, London, W2 1NY, UK; Wessex Regional Genomics Laboratory, Salisbury NHS Foundation Trust, Salisbury, SP2 8BJ, UK; University of Sussex, Brighton BN1 9RH, UK; Genomics England, Charterhouse Square, London, EC1M 6BQ, UK; Center for Genomic Medicine, Massachusetts General Hospital, Boston, MA 02114, USA; Genomics England, London, UK; William Harvey Research Institute, Queen Mary University of London, London, EC1M 6BQ, UK

## Abstract

The 100,000 Genomes Project (100KGP) diagnosed a quarter of recruited affected participants, but 26% of diagnoses were in genes not on the chosen gene panel(s); with many being *de novo* variants of high impact. However, assessing biallelic variants without a gene panel is challenging, due to the number of variants requiring scrutiny. We sought to identify potential missed biallelic diagnoses independent of the gene panel applied using GenePy - a whole gene pathogenicity metric.

GenePy scores all variants called in a given individual, incorporating allele frequency, zygosity, and a user-defined deleterious metric (CADD v1.6 applied herein). GenePy then combines all variant scores for individual genes, generating an aggregate score per gene, per participant. We calculated GenePy scores for 2862 recessive disease genes in 78,216 individuals in 100KGP. For each gene, we ranked participant GenePy scores for that gene, and scrutinised affected individuals without a diagnosis whose scores ranked amongst the top-5 for each gene. We assessed these participants’ phenotypes for overlap with the disease gene associated phenotype for which they were highly ranked. Where phenotypes overlapped, we extracted rare variants in the gene of interest and applied phase, ClinVar and ACMG classification looking for putative causal biallelic variants.

3184 affected individuals without a molecular diagnosis had a top-5 ranked GenePy gene score and 682/3184 (21%) had phenotypes overlapping with one of the top-ranking genes. After removing 13 withdrawn participants, in 122/669 (18%) of the phenotype-matched cases, we identified a putative missed diagnosis in a top-ranked gene supported by phasing, ClinVar and ACMG classification. A further 334/669 (50%) of cases have a possible missed diagnosis but require functional validation. Applying GenePy at scale has identified potential diagnoses for 456/3183 (14%) of undiagnosed participants who had a top-5 ranked GenePy score in a recessive disease gene, whilst adding only 1.2 additional variants (per individual) for assessment.

## Introduction

The 100,000 Genomes Project was a UK government funded research project led by Genomics England (GEL) to sequence 100,000 whole genomes for families predominantly presenting with rare disease.^1^ The project utilised a phenotype to genotype approach, whereby genome sequencing data were filtered using a pre-selected PanelApp^2^ gene panel or panels chosen by Genomics England based on the Human Phenotype Ontology (HPO)^3^ terms recorded at recruitment.^1; 4^ The project was completed in 2020 and yielded an overall diagnostic rate of ∽25% across all rare disease categories.^1; 5^ However, as ever-increasing numbers of researchers gained access to anonymised whole genome sequencing data from the 100,000 Genomes Project, additional diagnoses were made using methods that extended variant analysis beyond gene panels across more coding and non-coding regions, which have subsequently been returned to participants.^4^ As of 2022, 26% of all diagnoses returned by the 100,000 Genomes Project were from diagnoses not on the pre-selected gene panel applied, with many being pathogenic *de novo* coding variants.^4; 5^ However, assessing other variants such as biallelic variants is more burdensome, particularly without the use of gene panels due to the sheer number of variants that require scrutiny. This is because many are inherited from unaffected relatives and are carried at non-trivial allele frequencies in population databases. Furthermore, biallelic variation is often hard to interpret especially for compound heterozygotes where one variant may be pathogenic, and another may be a copy number variant, non-coding variant, or other variant of uncertain significance. This is where gene panels show their greatest utility since they can help narrow down variants to clinically relevant genes.^2^ However, this approach must be balanced against the potential of missing diagnoses outside of the original gene panel applied.

We sought to identify potential missed biallelic diagnoses in recessive disease genes independently of the gene panel applied using a whole genome pathogenicity metric called GenePy, pronounced “Jenni-pea 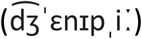”. GenePy (https://github.com/UoS-HGIG/GenePy-1.3) is a gene pathogenicity prioritization tool developed at the University of Southampton that transforms the interpretation of next generation sequencing data from the variant level to the gene or pathway level.^6^ GenePy incorporates allele frequency, individual zygosity (where a heterozygote scores one point and a homozygote scores two points), and a user-defined deleterious metric (such as CADD^7^) into a single variant score.

GenePy is defined as:

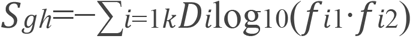

[Where h = individual; g = gene; k = variants; i = locus; *D*_*i*_ = allele deleteriousness; *f*_*i*_ = allele frequency; *f*_*i*1_ = allele 1; *f*_*i*1_ = allele 2]

GenePy then aggregates variant scores across genes in an additive manner, generating a single score, per gene, per individual that is represented in a GenePy matrix table (**Figure 1**). However, for large genes and intronic regions there is a potential to accumulate noise from low scoring variants. To mitigate this, GenePy can be customised to filter variants with high *in silico* scores only e.g. CADD score above a particular threshold. Additionally, GenePy can be applied across any defined interval and variant scores do not have to be summed across genes, e.g. one may choose to sum variants across a particular biological pathway or genomic region.

**Figure 1.**
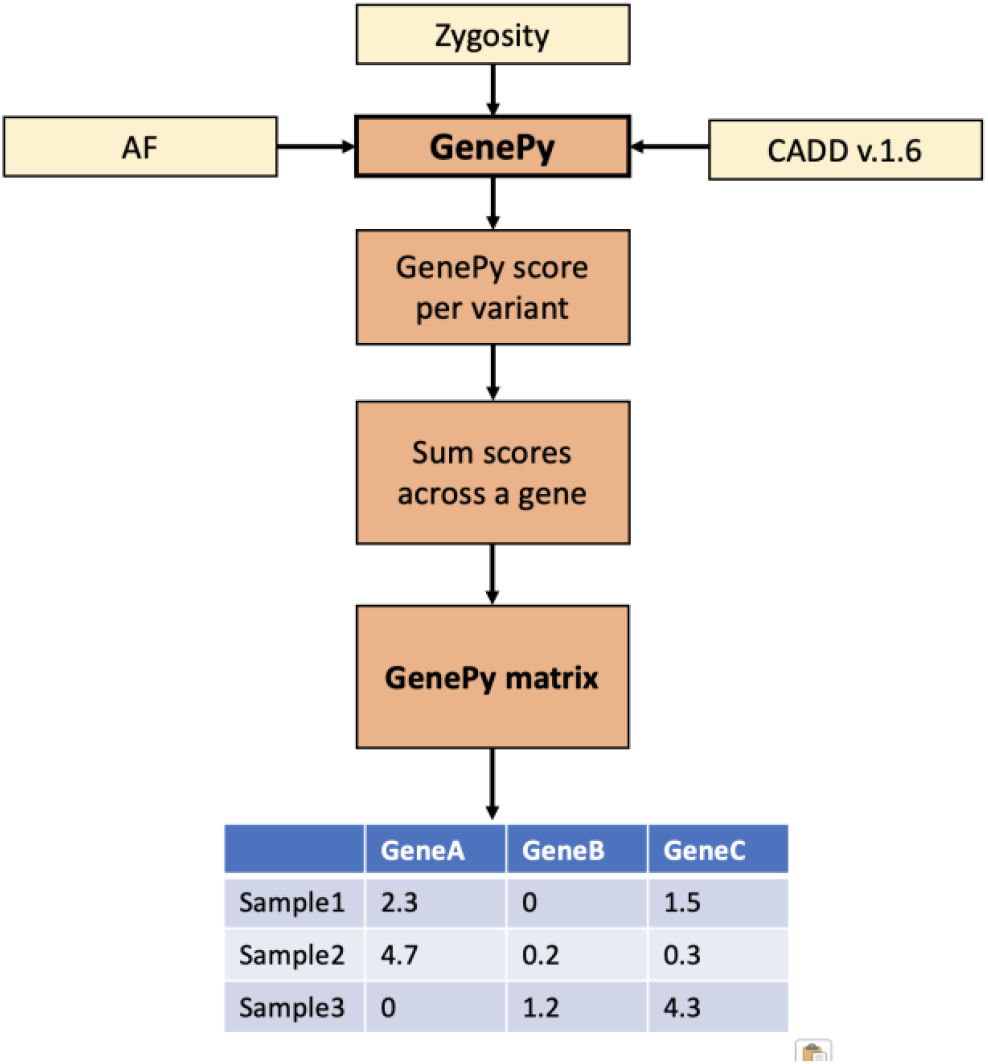
Overview of GenePy pathogenicity software and output. GenePy takes input from an annotated variant call file and uses allele frequency (such as defined by gnomAD), allele zygosity, and a deleterious metric, such as CADD. The GenePy software then scores each variant according to the GenePy equation. GenePy scores are summed per gene (although the user can also specify per exon, or per gene pathway if required). The resultant GenePy score (summed across a gene/region) is then represented in a GenePy matrix, with samples along the Y axis and gene/region along the X axis.

Upon generation of a GenePy matrix, GenePy scores can be compared across individuals in a cohort; GenePy scores are intuitive in that higher GenePy scores correlate with higher pathogenic variant burden such that individuals can be ranked for their score for any given gene, relative to all individuals with comparable input genomic data. GenePy scores are not easily compared between genes, without normalisation and adjustment for gene length. Even then, genes with alternative tolerance to dysfunctional variation are likely to exhibit very different GenePy score profiles. Instead, GenePy demonstrates the greatest utility when individual gene scores are compared across large numbers of individuals. Since GenePy is an additive score, individuals in large cohorts with the highest ranked GenePy scores will be enriched for biallelic disease. Given the potential for missed biallelic diagnoses in the 100,000 Genomes Project, we applied GenePy at scale in a panel-agnostic way to uplift diagnostic rates.

## Methods

### Access to 100,000 Genomes Project Data

Participants were recruited to the 100,000 Genomes Project with written consent. The full protocol is available here: https://doi.org/10.6084/m9.figshare.4530893.v7. Deidentified data from the project held are in the secure Genomics England Research Environment (RE).

We obtained access to 100,000 Genomes Project data following governance training and through membership of the ‘*Quantative Methods, Machine Learning, and Functional Genomics’* Genomics England Clinical Interpretation Partnership. We had an approved Genomics England Project (RR359).

In 2022, we accessed 78,216 whole genomes from affected and unaffected participants recruited to the 100,000 Genomes Project. We extracted participants’ affection status and any HPO terms associated with participants’ records. Using the package LabKey in R, we queried the ‘GMC Exit Questionnaire’ SQL table and extracted any diagnostic (likely pathogenic/pathogenic) variants returned to participants by the project.

### Curating a list of recessive disease genes

To target our method towards potential missed biallelic diagnoses, we curated a list of 2862 recessive disease genes using the OMIM^8^ database (downloaded in May 2022) and cross checked these findings with the GenCC database, whereby discrepancies in inheritance were examined more carefully.^9^ We then generated a bed file of gene coordinates for GRCh38 using the UCSC Genome Browser. The full gene list is available in Supplementary A.

### Application of GenePy

Within the Genomics England RE we applied GenePy v.1.3 (https://github.com/UoS-HGIG/GenePy-1.3) software to 78,216 participants in the 100,000 Genomes Project using CADD^7^ v1.6 as our deleterious metric and the gnomAD v.2.1.1 and V3^10^ databases as our reference for allele frequency. We selected variants with a minimum depth of 10, minimum GQ of 20, and mean GQ > 35 using vcftools. We applied a call-rate filter, whereby each variant had to be genotyped in at least 70% of the cohort. For downstream analysis, we only modelled and scored participant variants annotated as coding +/- 8 base pairs (on any transcript) and with a CADD score ≥15. We specified CADD as our input metric because it scores the greatest variety and number of variant types. We generated GenePy scores for 2862 recessive disease genes to create a matrix comprising GenePy scores for 2862 genes across 78,216 individuals. Of note, in addition to ‘affected’ participants, this cohort included many ‘control’ type individuals that represented unaffected parents of affected children and germline genomes of cancer patients.

For each of the 2862 recessive genes, we ranked every Genomic England participant’s GenePy score relative to one another e.g. the person with the highest GenePy score for *CFTR* would be ranked 1, and the person with the lowest GenePy score in *CFTR* would be ranked 78,216. After ranking, we arbitrarily assessed only individuals who ranked amongst the top-5 GenePy score for each gene. If two individual had identical scores, we retained all participants with a rank of 5 or less. We then removed any individuals who were coded as unaffected and affected individuals with insufficient phenotypic data in the form of HPO terms recorded. We next separated affected cases into those with a confirmed diagnosis returned by the 100,000 Genomes Project and those with a negative result. If the participant had a diagnosis returned, we assessed whether the established diagnostic variant was in a top-5 ranked GenePy score (**Figure 2**).

**Figure 2.**
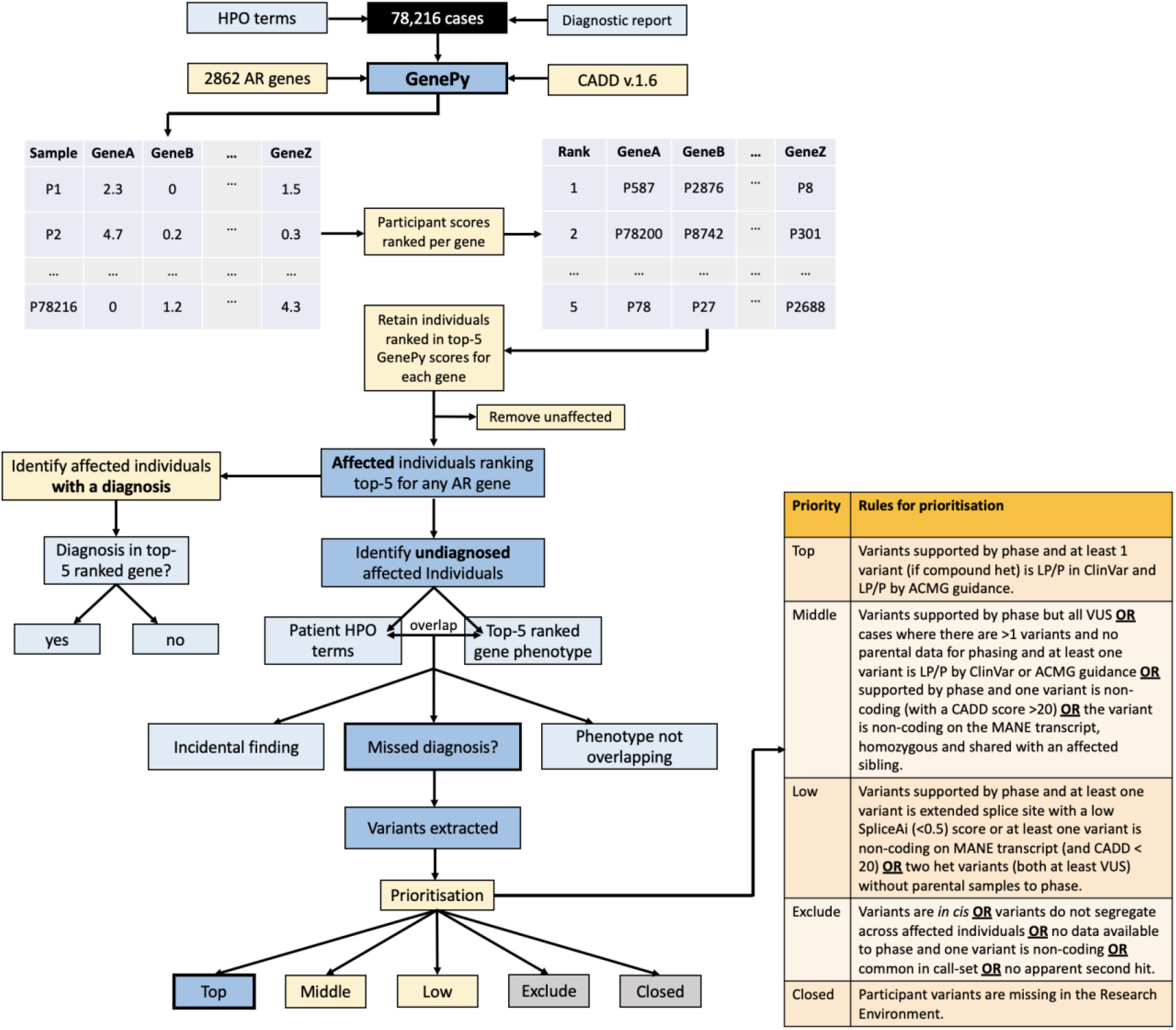
Workflow of GenePy applied to 78,216 cases in the 100,000 Genomes Project. GenePy scores were created for 2862 autosomal recessive genes in 78,216 participants, using CADD v.1.6 and gnomAD v.2.1.1. Participants scores were ranked across the cohort per gene, whereby those who ranked in the top 5 GenePy score for each gene were retained for downstream analysis. Unaffected individuals were removed. HPO terms from unaffected individuals without a diagnosis returned by the 100,000 Genomes Project were compared with the clinical features described for the autosomal recessive gene that the participant scored in the top 5 for. If the participant’s HPO terms overlapped with the gene that the person ranked in the top 5 for, we extracted the individual participant variants and assessed phase, ClinVar status, and applied ACMG guidelines. We then prioritised the findings according to the prioritisation rules, with ‘Top’ priority. being putative missed diagnoses, ‘Middle’ and ‘Low’ priority being of interest but lacking sufficient evidence, ‘Exclude’ being not diagnostic and ‘Closed’ being when the participants had been withdrawn from the Project.

For affected participants with a negative genome result, we extracted HPO terms from R Labkey and compared these HPO terms with the clinical features associated with the disease gene for which they scored in the top 5 rank. For example, if the participant had the HPO terms ‘pancreatic insufficiency’, ‘failure to thrive’ and ‘recurrent chest infections’ and they ranked third/3^rd^ for CFTR, we would compare their HPO terms with the clinical features of cystic fibrosis. This process was completed manually by a clinician who used clinical acumen, phenotypic descriptions and HPO terms listed in OMIM, and the clinical literature to help assess phenotype overlap. If the participant’s HPO terms were consistent with those for a gene that the same participant was ranked in the top 5 GenePy scores for (e.g. the participant had pancreatic insufficiency and recurrent chest infections and was ranked 3 in *CFTR)*, this was considered a potential missed diagnosis. If the disease-gene phenotype was unrelated to the participant’s clinical phenotype but represented a gene in the ACMG 78^11^ list or may represent an adult onset disease, this was considered a potential incidental finding. For these, we contacted the recruiting clinician to discuss the findings. If there was no correlation between the participant’s HPO terms and the clinical phenotype for the implicated disease gene, this was considered to be lacking phenotypic overlap and excluded from further consideration.

### Assessing potential missed diagnoses

When the participant’s phenotype was overlapping with the disease gene for which the participant ranked in the top 5, we extracted all variants from the participant’s variant call file with a CADD score ≥ 15. These variants were then prioritised by likelihood of being a missed biallelic diagnosis, taking into consideration variant phase where possible, ClinVar^12^ status, and variant curation by ACMG/AMP^13^ guidelines (**Figure 2**). Variants that were prioritised as ‘Top’ priority were considered putative missed diagnoses and mostly represented homozygous likely pathogenic/pathogenic variants or likely pathogenic/pathogenic compound heterozygous variants.

## Results

We applied GenePy to 2862 recessive disease genes in 78,216 cases recruited to the 100,000 Genomes Project. A summary of results is provided in **Figure 3**. For each gene we selected the top 5 ranked participants by GenePy score, which yielded a total of 9,404 unique participants, with some participants ranking top 5 for more than one recessive gene. 4713/9404 (50.1%) of the top ranked participants were unaffected participants, whereby anyone coded as unaffected (rare disease or cancer germline) represented 45% of the entire cohort. 4691/9404 (49.9%) were affected participants. Of the 4691 affected participants, 847/4691 (18.1%) already had a diagnosis returned by the 100,000 Genomes Project up to 2022. Of these, 599/847 (70.7%) had diagnoses in one of the top 5 ranked genes. 248/847 (29.3%) individuals had a diagnosis returned by GEL in an alternative gene. Of these, 87 individuals had a *de novo* pathogenic variant and 161 had a pathogenic variant in a dominant gene (either inherited from an affected individual or the participant was a from a singleton family).

**Figure 3.**
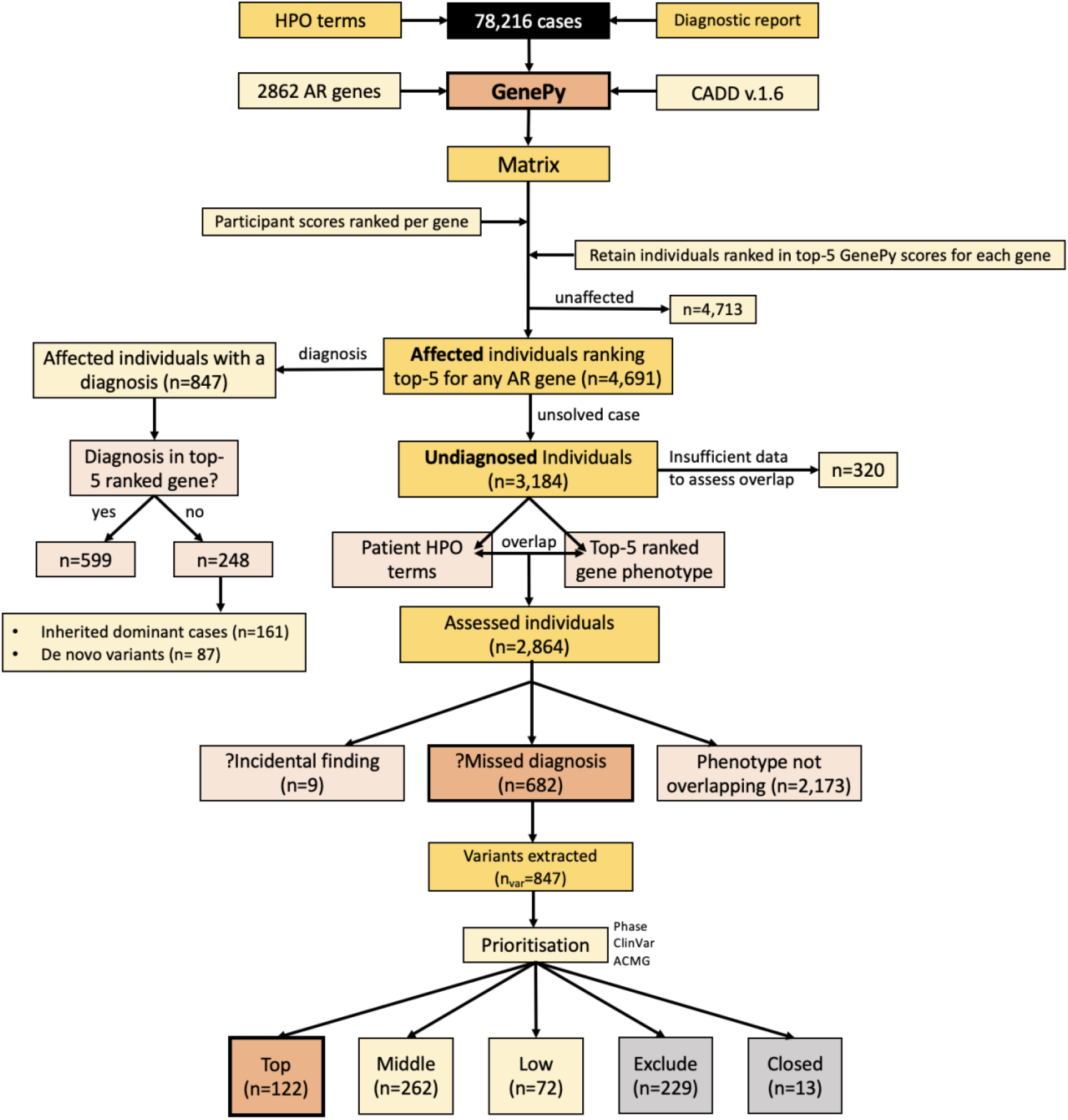
Summary of results. Results of GenePy applied to 2862 autosomal recessive disease genes in 78,216 individuals.

In total, there were 3184 affected individuals who had a ‘no diagnosis’ genome report returned by the 100,000 Genomes Project who were ranked in the top 5 GenePy scores for the 2862 computed recessive disease genes. For these cases, we compared the participant’s reported HPO terms with the clinical phenotype of the GenePy disease gene implicated in the participant. For 320 participants, there was insufficient HPO terms recorded to assess for phenotypic overlap between the participant’s clinical phenotype and that of the implicated disease gene; thus, these individuals were removed from downstream analysis. 2864 individuals had sufficient HPO terms to assess phenotype overlap and for 682/2864 (23.8%) of these cases, the participant’s HPO terms overlapped with the clinical presentation associated with the top 5 ranked GenePy disease gene. For 2173/2864 (75.9%) of cases, the phenotypes were non-overlapping and for 9/2864 (0.3%) of cases the phenotypes were not overlapping but the implicated gene was one of the ACMG 78 incidental finding genes.

For the 682 participants with a potential missed diagnosis, we extracted variants in their top 5-ranked gene with a CADD score ≥ 15 directly from their variant call file. In total we extracted 847 unique variants. Following prioritisation (**Figure 2**), we identified 122 top priority, putative missed diagnoses supported by phase, ClinVar^12^ classifications and ACMG/AMP guidelines.^13^ 262 individuals were assigned ‘Middle’ priority demonstrating supportive evidence for a potential missed diagnosis, whereby for many there was lack of phased data limiting diagnostic potential. 72 individuals had some, but weak evidence for a potential missed diagnosis for example due to one variant being non-coding on the matched annotation from ECBI and EMBL-EBI (MANE)^14^ transcript and were assigned ‘Low’ priority. 229 cases were ruled as non-diagnostic, typically due to the variants being *in cis*, being non-coding on the MANE transcript, not segregating with affected and related individuals, and being common in the 100,000 Genomes call-set (**Table 1**). In 13 cases, no variants were extracted because the individual had withdrawn from the 100,000 Genomes Project.

**Table 1.**
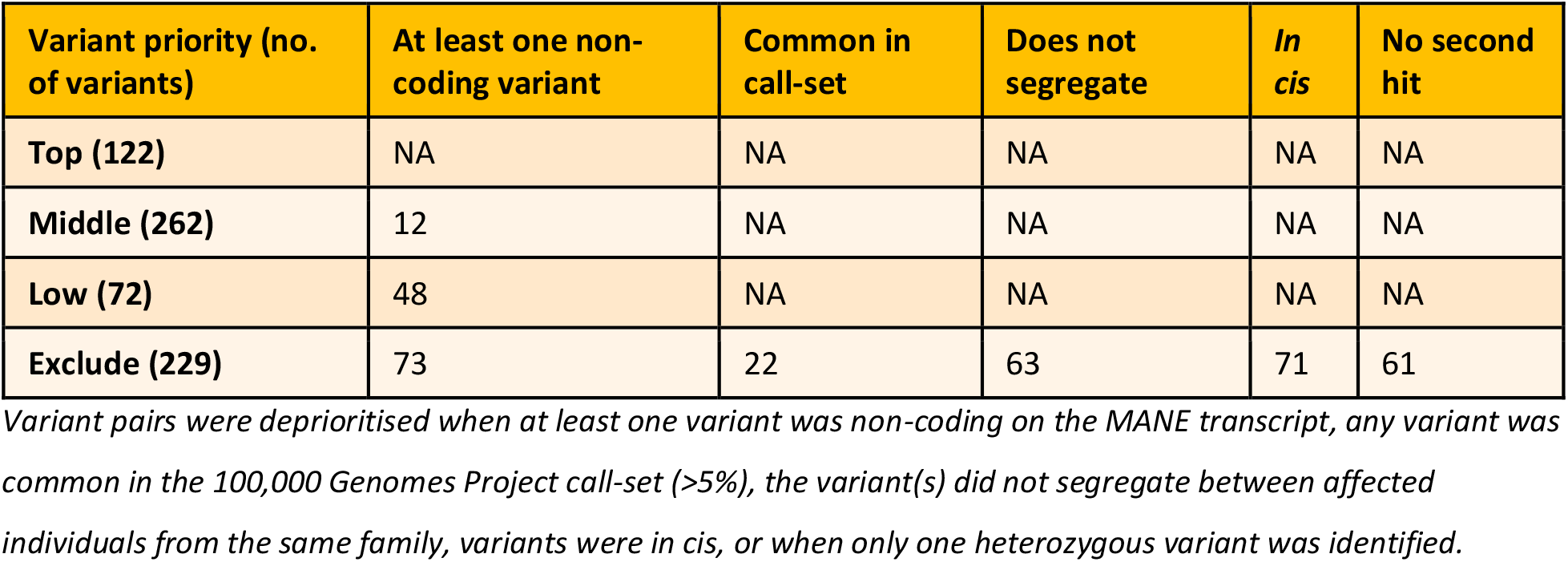
Flags applied to de-prioritise variants.

## Discussion

We applied a gene pathogenicity score, GenePy, to a cohort of 78,216 individuals recruited to the 100,000 Genomes Project. Utilising ranked individuals’ GenePy scores for 2862 recessive disease genes, we identified outliers with the highest GenePy scores per gene. We selected individuals who ranked in the top 5 scores for each gene, with an expectation that these individuals may harbour missed biallelic diagnoses.

847 individuals with a top 5 ranked GenePy score had a diagnosis returned by the 100,000 Genomes Project. 599/847 (71%) of these individuals had a diagnosis in a top 5 ranked gene, demonstrating how GenePy was able to rapidly recover 71% of diagnoses, showing potential diagnostic utility for both known and novel disease genes. The remaining 248 cases had diagnoses in dominant genes, with 81 diagnoses being *de novo* and 161 being inherited from an affected individual or the individual represented a singleton.

In total we identified 2864 undiagnosed individuals with top 5 ranked GenePy scores, of which 682/2864 (24%) had phenotypes overlapping with the clinical features of their top ranked recessive disease gene. Following prioritisation and removing 13 cases whereby participants had withdrawn from the 100KGP, 122/669 (18%) of the phenotype-matched cases had a putative missed diagnosis supported by phase, ClinVar classifications and ACMG/AMP guidelines. All these findings have since been returned to Genomics England through their Diagnostic Discovery Pathway. For 334/669 (50%) of individuals, we identified variants of interest in a disease gene consistent with the participant’s phenotype with some supportive evidence for pathogenicity, but often phase could not be determined due to missing parental data. Additionally, for many of these cases, the variants contributing to the high GenePy scores were classified as VUSs and therefore require additional functional work-up. These variants are being reviewed by a clinical scientist in an NHS accredited diagnostic laboratory. Whilst follow-up of these variants is outside the scope of this research project, many of these variants, even those prioritised in the low category, may represent pathogenic variants. For example, non-coding variants were assigned to a lower priority grouping, despite them having a CADD score ≥ 15. It is hoped that many of these variants may be functionally investigated in the future as high-throughput methods to model VUSs advance.

In total, GenePy has identified potential missed diagnoses in 456/2864 (16%) of undiagnosed individuals who had a top-5 ranked GenePy score in a recessive disease gene. On average this resulted in the curation of 1.2 additional variants per participant. Therefore, the application of GenePy successfully uplifted diagnosis rates without adding large variant numbers requiring time-consuming manual curation for diagnostic laboratories to assess and classify.

GenePy^6^ is an open-source transferrable piece of software that can be successfully applied at scale. GenePy matrices can be used as reference datasets for other cohorts applying the same GenePy methods i.e. when applying the same deleterious metric, population reference database and quality control thresholds. For example, GenePy may be applied to a cohort of 10 samples, whereby these 10 individuals’ GenePy scores could be ranked against a larger GenePy matrix comprising 100,000 individuals. However, GenePy matrices for genome sequencing data should only be compared with other genome sequencing datasets, unless restricted to the same target regions of exome data.

### Limitations and opportunities

The application of GenePy to the 100,000 Genomes Project is not without its limitations. For one, we used an entirely arbitrary cut off of 5 when we ranked individuals. It is entirely possible that a more permissive value may capture a wider range of diagnoses; however, this must be balanced with the additional number of variants, per individual, that would require further scrutiny by clinical laboratories.

We assessed for phenotype overlap between the participants’ HPO terms and the clinical features described for the disease-gene in which the participants ranked in the top 5 GenePy scores. For 320 cases, the HPO terms were so limited that it was not possible to assess overlap.

This represents a real-world limitation of sequencing studies whereby there is often variability in how submitters record phenotype data and highlights the importance of accurate phenotyping. This phenotype comparison step was performed manually on 2864 cases. Application of automated methods to compare participant HPO terms with disease gene phenotypes may prove more time efficient for GenePy applied at scale, however it is unlikely that clinical or diagnostic laboratories applying GenePy would be reviewing thousands of individuals at once, but rather on a case-by-case basis. Additionally, automated methods lack the clinical knowledge and experience of a clinician or clinical scientist that may be better able to intelligently compare groups of similar phenotypes.

In our application of GenePy we used CADD v.1.6 to capture and model the greatest breadth of variation in an unbiased way, but it may be that incorporation of other metrics for different variant types (e.g. REVEL^15^ for missense) may prove more sophisticated in an improved model. However, this is likely to require machine learning to apportion *in silico* weightings fairly for different variant types. We also applied a CADD cut off of ≥ 15 to avoid individuals accruing high GenePy scores in genes of increasing length, where there was a higher chance of finding multiple ultrarare variants by pure chance that would score highly in GenePy. Whilst we are confident that using CADD ≥ 15 reduced a lot of noise and helped isolate pathogenic variants, we accept that this approach risks missing some pathogenic variants with lower CADD scores.

GenePy currently does not utilise phased data, meaning that some high scores may represent variants inherited *in cis*; indeed, we observed this in 71 cases (**Table 1**). However, we were conscious not to limit GenePy to nuclear families with parental data since this does not represent a real-world example and would disadvantage non-parent/child families, where phase cannot be determined. In the future, this could perhaps be mitigated with long read sequencing data.

Whilst we applied GenePy herein focusing on identification of potential missed recessive disease, there may also be opportunities to apply it in autosomal dominant diseases. When we scrutinised the variants of individuals with potential missed diagnoses, we identified 61 individuals that ranked in the top 5 GenePy scores for a given gene, yet they only had one variant with a CADD score ≥ 15 in that gene. Most commonly these individuals harboured predicted loss-of-function variants which are upweighted in the GenePy statistic. Therefore, there may be utility of GenePy in haploinsufficient disease genes, but it is likely that a more stringent CADD cut off, such as ≥ 20, or limiting the GenePy statistic to the highest scoring variant is necessary to apportion lower GenePy scores to individuals who would otherwise accrue high scores from multiple rare, but benign variants with lower CADD scores.

GenePy also has potential to identify novel disease genes. If multiple top-ranking individuals across the same novel gene share similar clinical features, this may support the discovery of new disease genes. For novel haploinsufficient genes, unpublished data from our research group suggest that GenePy performs best when limited to high CADD scores e.g. CADD >20, whereas recessive genes may benefit from a more permissive CADD cut off.

## Conclusion

The application of GenePy to ∽78,000 individuals in the 100,000 Genomes Project has identified 122 putative missed biallelic diagnoses in known autosomal recessive disease genes that are being returned to participants through the Genomics England diagnostic discovery pathway. Selecting the top 5 ranked individuals for 2864 autosomal recessive genes yielded review of only 1.2 additional variants per individual, rendering GenePy a useful tool to identify biallelic variants of interest without significantly burdening diagnostic laboratories with additional variants to assess. A dilemma for many diagnostic laboratories is how to limit number of variants requiring assessment without missing diagnoses. Whilst strategies to prioritise dominant diseases are well established e.g. de novo analysis or Exomiser^16^, there are limited tools for prioritising recessive conditions. We attest that GenePy is a useful panel-agnostic adjunct to exome and genome analysis pipelines to uplift diagnoses of recessive disease.

## Data Availability

Access to the 100KGP dataset analysed in this study is only available as a registered GeCIP member in the Genomics England Research Environment, but restrictions apply to the availability of these data due to data protection and are not publicly available. Information regarding how to apply for data access is available at the following url: https://www.genomicsengland.co.uk/about-gecip/for-gecip-members/data-and-data-access/. All data shared in this manuscript were approved for export by Genomics England. The datasets and code supporting the current study are fully accessible within the Genomics England Research Environment.

## Acknowledgements

This research was made possible through access to the data and findings generated by the 100,000 Genomes Project. The 100,000 Genomes Project is managed by Genomics England Limited (a wholly owned company of the Department of Health and Social Care). The 100,000 Genomes Project is funded by the National Institute for Health Research and NHS England. The Wellcome Trust, Cancer Research UK and the Medical Research Council have also funded research infrastructure. The 100,000 Genomes Project uses data provided by patients and collected by the National Health Service as part of their care and support. We would further like to extend our thanks to all the patients and their families for participation in the 100,000 Genomes Project.

## References

1. 100, G.P.P.I. (2021). 100,000 Genomes Pilot on Rare-Disease Diagnosis in Health Care— Preliminary Report. New England Journal of Medicine 385, 1868–1880.

2. Martin, A.R., Williams, E., Foulger, R.E., Leigh, S., Daugherty, L.C., Niblock, O., Leong, I.U., Smith, K.R., Gerasimenko, O., and Haraldsdottir, E. (2019). PanelApp crowdsources expert knowledge to establish consensus diagnostic gene panels. Nature genetics 51, 1560–1565.

3. Robinson, P.N., Köhler, S., Bauer, S., Seelow, D., Horn, D., and Mundlos, S. (2008). The Human Phenotype Ontology: a tool for annotating and analyzing human hereditary disease. The American Journal of Human Genetics 83, 610–615.

4. Seaby, E.G., Thomas, N.S., Webb, A., Brittain, H., Taylor Tavares, A.L., Baralle, D., Rehm, H.L., O’Donnell-Luria, A., and Ennis, S. (2022). Targeting de novo loss-of-function variants in constrained disease genes improves diagnostic rates in the 100,000 Genomes Project. Hum Genet, 1–12.

5. Rehm, H.L. (2022). Time to make rare disease diagnosis accessible to all. Nature Medicine 28, 241–242.

6. Mossotto, E., Ashton, J.J., O’Gorman, L., Pengelly, R.J., Beattie, R.M., MacArthur, B.D., and Ennis, S. (2019). GenePy - a score for estimating gene pathogenicity in individuals using next-generation sequencing data. BMC Bioinformatics 20, 254.

7. Kircher, M., Witten, D.M., Jain, P., O’Roak, B.J., Cooper, G.M., and Shendure, J. (2014). A general framework for estimating the relative pathogenicity of human genetic variants. Nat Genet 46, 310–315.

8. Amberger, J.S., Bocchini, C.A., Schiettecatte, F., Scott, A.F., and Hamosh, A. (2015). OMIM. org: Online Mendelian Inheritance in Man (OMIM®), an online catalog of human genes and genetic disorders. Nucleic acids research 43, D789–D798.

9. DiStefano, M.T., Goehringer, S., Babb, L., Alkuraya, F.S., Amberger, J., Amin, M., Austin-Tse, C., Balzotti, M., Berg, J.S., Birney, E., et al. (2022). The Gene Curation Coalition: A global effort to harmonize gene-disease evidence resources. Genet Med.

10. Karczewski, K.J., Francioli, L.C., Tiao, G., Cummings, B.B., Alföldi, J., Wang, Q., Collins, R.L., Laricchia, K.M., Ganna, A., Birnbaum, D.P., et al. (2020). The mutational constraint spectrum quantified from variation in 141,456 humans. Nature 581, 434–443.

11. Gambin, T., Jhangiani, S.N., Below, J.E., Campbell, I.M., Wiszniewski, W., Muzny, D.M., Staples, J., Morrison, A.C., Bainbridge, M.N., Penney, S., et al. (2015). Secondary findings and carrier test frequencies in a large multiethnic sample. Genome Med 7, 54.

12. Landrum, M.J., Lee, J.M., Riley, G.R., Jang, W., Rubinstein, W.S., Church, D.M., and Maglott, D.R. (2014). ClinVar: public archive of relationships among sequence variation and human phenotype. Nucleic acids research 42, D980–D985.

13. Richards, S., Aziz, N., Bale, S., Bick, D., Das, S., Gastier-Foster, J., Grody, W.W., Hegde, M., Lyon, E., Spector, E., et al. (2015). Standards and guidelines for the interpretation of sequence variants: a joint consensus recommendation of the American College of Medical Genetics and Genomics and the Association for Molecular Pathology. Genet Med 17, 405–424.

14. Morales, J., Pujar, S., Loveland, J.E., Astashyn, A., Bennett, R., Berry, A., Cox, E., Davidson, C., Ermolaeva, O., and Farrell, C.M. (2022). A joint NCBI and EMBL-EBI transcript set for clinical genomics and research. Nature, 1–6.

15. Ioannidis, N.M., Rothstein, J.H., Pejaver, V., Middha, S., McDonnell, S.K., Baheti, S., Musolf, A., Li, Q., Holzinger, E., and Karyadi, D. (2016). REVEL: an ensemble method for predicting the pathogenicity of rare missense variants. The American Journal of Human Genetics 99, 877–885.

16. Smedley, D., Jacobsen, J.O., Jäger, M., Köhler, S., Holtgrewe, M., Schubach, M., Siragusa, E., Zemojtel, T., Buske, O.J., and Washington, N.L. (2015). Next-generation diagnostics and disease-gene discovery with the Exomiser. Nature protocols 10, 2004–2015.

